# Awareness of fifth metatarsal stress fractures among soccer coaches in Japan: A cross-sectional study

**DOI:** 10.1101/2022.11.01.22281790

**Authors:** Takayuki Miyamori, Masafumi Yoshimura

## Abstract

Although a fifth metatarsal stress fracture is the most frequent stress fracture in soccer players, knowledge of fifth metatarsal stress fractures among soccer coaches is unclear. Therefore, we performed an online survey of soccer coaches affiliated with the Japan Football Association to assess their knowledge of fifth metatarsal stress fractures. A total of 117 soccer coaches participated in an original online survey. Data on age, sex, types of coaching license, coaching category, types of training surface, awareness of fifth metatarsal stress fractures, and measures employed to prevent fifth metatarsal stress fractures were collected using the survey. A total of 87/117 coaches were aware of fifth metatarsal stress fractures; however, only 30% reported awareness of preventive and treatment measures for fifth metatarsal stress fractures. Licensed coaches (i.e., licensed higher than level C) were also more likely to be aware of fifth metatarsal stress fractures than were unlicensed coaches. Furthermore, although playing on artificial turf is an established risk factor for numerous sports injuries, soccer coaches who usually trained on artificial turf were more likely to be unaware of the risks associated with fifth metatarsal stress fractures than were coaches who trained on other surfaces (e.g., clay fields). Soccer coaches in the study population were generally aware of fifth metatarsal stress fractures; however, most of them were not aware of specific treatment or preventive training strategies for fifth metatarsal stress fractures. Additionally, coaches who practiced on artificial turf were not well educated on fifth metatarsal stress fractures. Our findings raise awareness of fifth metatarsal stress fractures to improve the education of soccer coaches regarding injury prevention strategies.

## Introduction

A fifth metatarsal stress fracture (MT-5) is the most frequent stress fracture in soccer players. In particular, zones II and III in the MT-5 Torg classification are located in the proximal shaft, and the incidence of stress fractures in these zones is also high; these fractures are known as Jones fractures [1,2]. The fifth metatarsal is prone to bone stress owing to poor blood supply and is commonly reinjured; thus, surgical treatment is often selected [3]. The estimated return-to-play time for soccer players with MT-5 is generally between 3 and 5 months; therefore, MT-5 is considered a career altering obstacle [3]. In injury surveys of European professional soccer players, the incidence of stress fractures was approximately 5% of all injuries, 78% of which were MT-5 [4].

Because of the high incidence and substantial return-to-play time of MT-5, team staff need to consider the risk factors for developing MT-5 during training. In previous studies, age and sex were found to be risk factors for MT-5 [5,6]. Anatomical studies have also reported immobilization of the lateral plantar fascia and short peroneus muscle involvement in induration of the tendon [2]. Nutritionally, a deficiency in 25-hydroxyvitamin D, an indicator of calcium metabolism, may likewise affect the risk of MT-5 [7]. In addition, kinematic problems, such as foot distraction tendencies during loading [8], limited range of motion of hip internal rotation [7], and toe grip muscle weakness [8], may lead to decreasing balance capacity in lower extremities; thus, increasing the planter pressure on the base of the fifth metatarsal.

During practice sessions, doctors and physical therapists are sometimes present at the training sites and they have opportunities to provide MT-5 prevention advice and training to athletes during regular training and games. However, coaches’ knowledge regarding MT-5 prevention is unknown. Therefore, we conducted a questionnaire survey of soccer coaches affiliated with the Japan Football Association (JFA) to investigate their knowledge of MT-5 and its prevention and treatment. We hypothesized that awareness of MT-5 among soccer coaches varies with the level of licensure and the coaching environment.

## Materials and methods

A cross sectional study was conducted. A total of 150 soccer coaches on the JFA official team were invited to participate in this study through the JFA Official Coaching Course at Juntendo University in Japan and direct email between 15^th^ of January and 30^th^ of March in 2020. A computer-based survey was created, and a research request was sent to potential respondents who provided consent. Each participant entered the mobile terminal using a personalized code. The exclusion criteria were failure to provide informed consent to complete the survey. Additionally, if more than one incomplete response in the questionnaire was confirmed, all the individual’s data was excluded from the analysis. The inclusion and exclusion criteria used are indicated in Fig 1.

**Fig 1.**
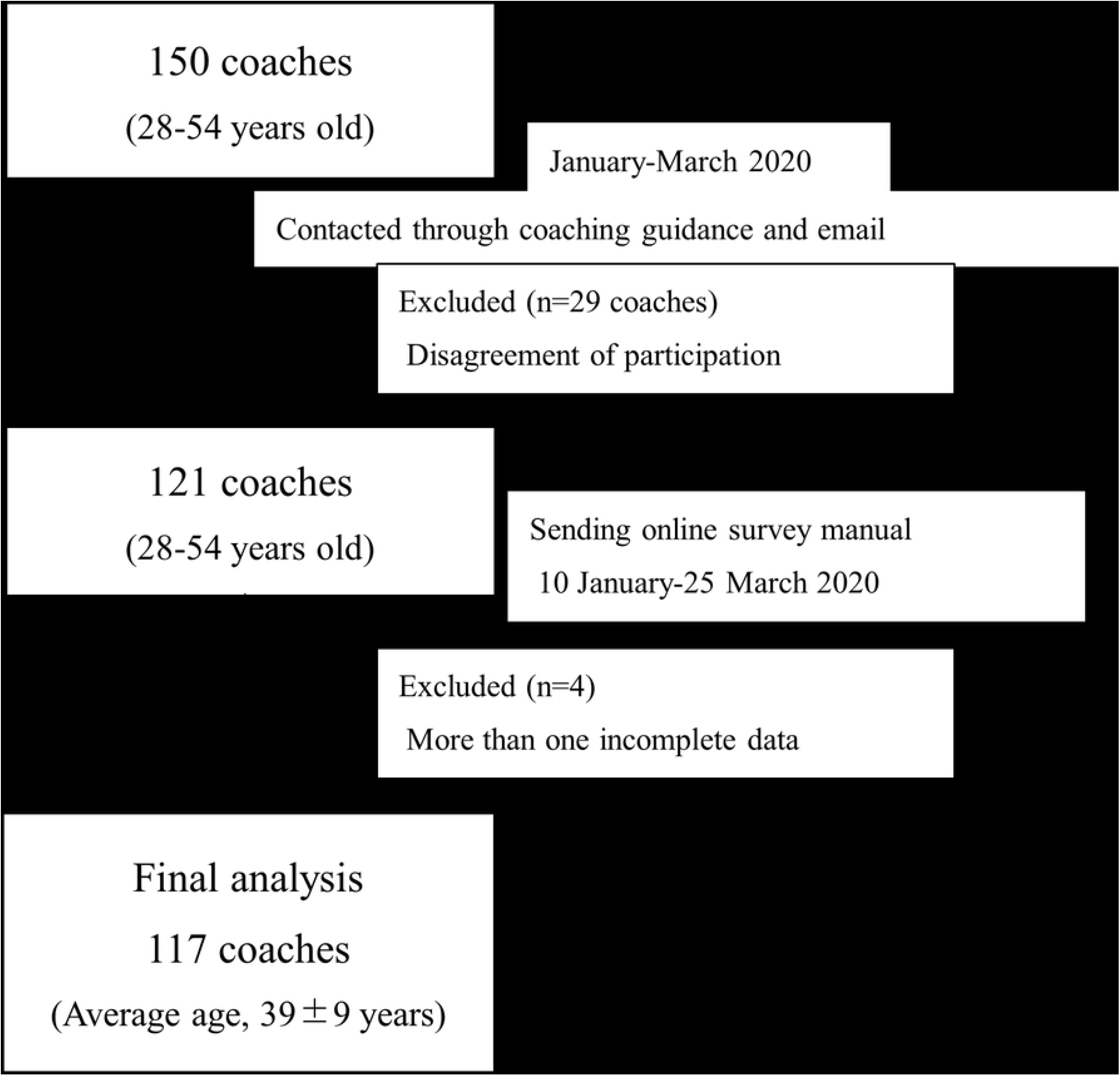
Flowchart of participant recruitment and screening.

The survey items included age, sex, years of playing soccer, years of coaching experience, level of JFA license (none, D, C, B, A, and S, which is the highest level of license), coaching category (elementary student–J league), type of training surface (artificial turf and other than artificial turf), training frequency per week, training hours per session, awareness of MT-5, and awareness of preventive training strategies for MT-5. In addition, 20 participants across all coaching categories were randomly assigned to complete the survey a second time 1 week after their first survey completion to identify the reliability of the self-reported questionnaire. Statistical analyses were performed using SPSS version 24 (IBM Corp., Armonk, NY). The number of respondents and overall proportion of responses for each question item in the survey were calculated. Univariate analysis was performed to examine the relationship between license type and MT-5 awareness. Additionally, the odds ratio (OR), 95% confidence interval, and *p* value of each license were calculated with “no license” as the reference value. Furthermore, a chi-square test was performed to examine the relationship between training surface types and awareness of MT-5. Statistical significance was set at *p* < 0.05.

The survey was anonymized to ensure privacy. A research ID corresponding to the participant’s name was created. Registration for the survey was also conducted using the research ID. To prevent unauthorized access, an encrypted passcode was set separately from the research ID, and responses could only be made by entering this passcode to ensure confidentiality. This study was approved by the International University of Health and Welfare Ethics Review Board (approval number: 16-Io-203). All survey respondents provided written informed consent.

## Results

We contacted 150 potential participants, and the data from 117 respondents (average age, 39 ± 9 years; effective response rate: 78%) were analyzed. Table 1 shows the number and proportion of responses for each question item in the survey. The coaching category with the highest frequency was high school, which accounted for 51.3% of the total. More than half (54.7%) of the respondents coached on artificial turf. Approximately 30% of respondents had an A-level license, with the fewest respondents having a D-level license. Of the 117 total respondents, 87 (74.4%) were aware of MT-5, although only 30% implemented treatment methods and preventive training strategies. MT-5 prevention methods were assessed using a free-format question; the most frequent response was adjusting the amount of training; other responses included trunk training, ankle tube training, and changing spike cleats. Table 2 shows the types of licenses and the results of the univariate analysis of the degree of MT-5 awareness. The univariate analysis showed that the OR of being unaware of MT-5 was lower for all licenses than it was for unlicensed coaches. Holding a D-level license reduced the odds of being unaware of MT-5 by 75%; however, this finding was not statistically significant (*p* = 0.30). Coaches with C-level, B-level, A-level, and S-level licenses were significantly more likely to be aware of MT-5 than were unlicensed coaches (all *p* < 0.05). Table 3 summarizes the results of the chi-square test for the type of training surface and the degree of MT-5 awareness. Coaches training on artificial turf were more likely to be unaware of MT-5 than were coaches training on other surface types (*p* < 0.05).

**Table 1.** Responses to the questionnaire.

|  | Possible responses | % | n |
| --- | --- | --- | --- |
| Coaching categories | Primary school | 7.7 | 9 |
|  | Junior high school | 17.1 | 20 |
|  | High school | 51.3 | 60 |
|  | University | 19.7 | 23 |
|  | Semi professional | 3.4 | 4 |
|  | Professional | 0.9 | 1 |
| Use of artificial turf | Yes | 54.7 | 64 |
| Certified license (level) | No license | 12.8 | 15 |
|  | D | 2.6 | 3 |
|  | C | 21.4 | 25 |
|  | B | 24.8 | 29 |
|  | A | 32.5 | 38 |
|  | S | 6 | 7 |
| Knowledge of MT-5 | Yes | 74.4 | 87 |
| **Knowledge of MT-5 treatment methods** | Yes | 29.9 | 35 |
| **Preventive training** | Yes | 30.8 | 36 |

MT-5, fifth metatarsal stress fracture; %, percentage; n, number of coaches

**Table 2.** Relationship between license level and MT-5 recognition.

|  |  | **OR** | **95% CI** | **p-value** |
| --- | --- | --- | --- | --- |
| **Certified license (level)** | ref. = no license | 0.35 |  | <0.05 |
|  | D | 0.25 | 0.02–3.47 | 0.3 |
|  | C | 0.1 | 0.02–0.43 | <0.05 |
|  | B | 0.16 | 0.04–0.63 | <0.05 |
|  | A | 0.11 | 0.03–0.44 | <0.05 |
|  | S | 0.08 | 0.01–0.90 | <0.05 |

OR, odds ratio; 95% CI, 95% confidence interval; ref., reference

**Table 3.** Relationship between type of training surface and MT-5 recognition.

|  |  | **Type of training surface\*** |  | **Total** |
| --- | --- | --- | --- | --- |
|  |  | **Artificial turf** | **Other than artificial turf** |  |
| **MT-5 knowledge** | Yes | 39 (60.9%) | 48 (90.6%) | 87 |
|  | No | 25 (39.1%) | 5 (9.4%) | 30 |
| **Total** |  | 64 | 53 | 117 |

\*Percentage of coaches

## Discussion

This was the first online questionnaire survey of soccer coaches affiliated with a JFA-certified team in which the license status of coaches, training surfaces, and MT-5 awareness were examined. Among the 117 participants surveyed, 87 (74.4%) of soccer coaches were familiar with MT-5. Licensed coaches (C, B, A, and S levels) were significantly more likely to be aware of MT-5 than were unlicensed coaches. However, only 30% of coaches implemented treatment and preventive training. Clearly, soccer coaches lack knowledge on how to prevent and treat MT-5.

A previous study surveying lower extremity joint injuries among female soccer players and instructors found that, although coaches were more aware of injuries than were athletes, they did not adequately understand the risk factors for injuries or specific prevention strategies [9]. In addition to tactical and technical training instruction, the JFA official coach license curriculum covers sports injury prevention and first aid. Furthermore, the JFA published a guide of soccer medicine for athletes and coaches in 2005 and advocated a method for evaluating the physical strength and conditioning necessary for soccer, as well as a method for managing typical orthopedic and medical diseases [10]. Lower limb and trunk injuries account for 90% of injuries in soccer, with particular attention given to Osgood-Schlatter disease and knee and ankle joint soft tissue injuries (meniscal and ligament injuries), along with lumbar injuries during the growth period in athletes [10]. However, currently, coaches do not adequately address factors related to MT-5 using preventive interventions and rehabilitation.

Approximately 50% of the respondents in the current study coached high school athletes; meanwhile, 54.7% of all coaches trained their athletes on artificial turf. Those who coached on artificial turf had lower awareness of MT-5 injuries than did those who coached on other surfaces. Since 2001, the use of long-pile artificial turf has increased rapidly in Japan [11]. Although artificial turf has many advantages, including reduced ground maintenance costs, epidemiological studies have shown that artificial turf causes an increased burden on the feet and knee joints, leading to greater injuries compared with other surfaces [12-14]. Ekstrand et al. [14] determined that training and playing matches are more frequently conducted on artificial grass than on natural grass in high school and college. Turning motions on artificial grass have been shown to cause rotational stress on the foot, and the accumulation of stress increases as the intensity of competition increases from high school to university. [13] In recent studies, players with a high body mass index also reported an increase in plantar load due to a decreased balance ability during play [15] and an increase in mechanical stress on the fifth metatarsal secondary to changes in lateral plantar pressure during turning [16]. Furthermore, Miyamori et al. [17] showed that training or playing on artificial turf increases the risk of developing MT-5 compared with clay field. Together, these studies showed that increased play on artificial turf may increase the risk of stress fractures of the lower extremities, such as MT-5.

To reduce the incidence of MT-5, doctors and physical therapists need to provide specialized training regarding medical science and preventive training methods for coaches. Future research should focus on determining other risk factors for lower extremity stress fractures, like MT-5, which tend to be specific to soccer players. It is necessary to develop a training program that considers the risk factors for MT-5, which would improve athletes’ self-management ability, including injury prevention. These approaches can strengthen individual athletes and teams.

This study had three main limitations and future directions that warrant discussion. First, our sample size was relatively small; however, we were able to describe different aspects across a range of age groups and categories. Second, some of the coaches who participated in our survey may have been medically trained to be aware of MT-5 and provide education regarding MT-5 prevention and therapy. This would relieve the pressure on the coaching staff by directing their efforts toward MT-5 awareness. Future studies must examine the composition of team staff to determine the status of the professional injury prevention system and improve athletes’ awareness of sports injury management. Finally, this study was limited to the Japanese soccer community and must be expanded to include coaches and medical staff across Asia, Europe, and worldwide. Sports injury surveillance systems are currently being developed for soccer [18]. In the future, it will be necessary to integrate and disseminate all injury reports from Japanese soccer in collaboration with the JFA Sports Medicine Committee and the Japan professional soccer league. This approach will make it possible to identify and share risk factors for sports injuries, including MT-5, which is a universal problem, ultimately contributing to sports injury prevention.

## Conclusion

We conducted an MT-5 awareness survey among soccer coaches. Coaches without a JFA-certified coaching license had lower awareness of MT-5 than did coaches with such licenses (C level and above). In contrast, although licensed coaches understood the nature of the injury, only a third understood and implemented treatment methods and preventive training. Additionally, even though playing on artificial turf is associated with a higher injury incidence, those who coached on artificial turf had lower awareness of MT-5 than did those who coached on other surfaces. In the future, JFA coach training sessions should increase coaches’ knowledge of MT-5 pathologies and suggest concrete preventive training methods to reduce the risk of developing MT-5.

## Data Availability

All relevant data are within the manuscript and its Supporting Information files.

## Acknowledgements

We would like to thank Ryuichi Sawa and Yu Shimasaki for their valuable help with this study. We would like to thank Editage (www.editage.com) for English language editing.

